# 4-Phenylbutyrate for STXBP1 and SLC6A1. Safety, tolerability, seizure, and EEG outcomes. A case series at 2 centers

**DOI:** 10.1101/2024.11.06.24316676

**Authors:** Zachary M Grinspan, Jacqueline Burré, Jennifer Cross, M Elizabeth Ross, Amelia Stone, Natasha Basma, Kerry Gao, Jing-Qiong Kang, Jaehyung Lim, Elizabeth Dubow, Maria Abila, Andrea Miele, Scott Demarest

## Abstract

**Introduction:** Pathogenic mutations in STXBP1 and SLC6A1 can cause developmental delay and epilepsy. 4-phenylbutyrate (4PB), a drug used for urea cycle disorders, rescues dysfunction in pre-clinical models for both genes, suggesting an opportunity for drug repurposing.

**Methods:** We conducted a single-treatment group, multiple-dose, open-label study of 4PB (as glycerol phenylbutyrate) in children with pathogenic STXBP1 and SLC6A1 mutations at two centers (NCT04937062). Enrolled participants were monitored for 4 weeks (baseline) then received 4PB for 10 weeks, with an option for extended use. Endpoints were safety and tolerability (primary) as well as seizure burden, EEG abnormalities, quality of life, development, behavior, and sleep (exploratory). We report safety, tolerability, EEG, and seizure outcomes.

**Results:** We enrolled 20 children (10 STXBP1, 10 SLC6A1; median age 5 years, absolute range 4 months to 11 years; 14 males; all White, 2 Hispanic). There were six serious adverse events, one attributed to 4PB (hospitalization for metabolic acidosis). Other common adverse events included a honey-like odor, sedation, and anorexia. After one participant initially withdrew for metabolic acidosis (SLC6A1), 18 of the 19 remaining participants opted for extended use. Of these 18, 17 (94%) continued 4PB within 10% of the target dose (11.2 mL/m^2/day) at the last clinical visit (2-3 years). At baseline, every child had abnormal EEG findings (seizures, paroxysmal or generalized slowing, or epileptiform discharges). For STXBP1, EEG improvements after one year of 4PB include (1) six with EEG seizures before 4PB versus two after and (2) nine with epileptiform discharges on initial EEGs versus four after. For SLC6A1, there was no clear pattern of EEG evolution. After 10 weeks of 4PB, seizures (or spells) were reduced in STXBP1 for six (60%), and in SLC6A1 for seven (70%). Seizure outcomes at the last visit were as follows. For STXBP1, three children were seizure-free and six had seizures daily to monthly; for SLC6A1, eight had sustained reduction in seizures, typically with seizure freedom and brief relapses that resolved after weight adjustment of the 4PB dose.

**Conclusion:** This case series found 4PB was safe and well-tolerated and reduced seizures for STXBP1 and SLC6A1 disorders.

**KEY POINTS:** *Question:* Is 4-phenylbutyrate (4PB; a drug used for urea cycle disorders) safe, tolerable, and effective for people with developmental delay and seizures due to mutations in STXBP1 or SLC6A1?

*Findings:* In a single-treatment group, multiple-dose, open-label study of 4PB at two centers, treatment of 20 children (10 STXBP1 and 10 SLC6A1) with 4PB (as glycerol phenylbutyrate) was safe (no new side effects) and well tolerated (19 enrolled in extended use). For both disorders, seizures improved in the short term (6 STXBP and 7 SLC6A1 with fewer seizures after 6 weeks) and long term (3 STXBP1 and 8 SLC6A1 with sustained seizure reduction after 2-3 years of use).

*Meaning:* 4PB is a safe and well-tolerated repurposed drug that may improve seizure control in STXBP1 and SLC6A1.

## INTRODUCTION

Pathogenic mutations in syntaxin-binding protein 1 (STXBP1) cause STXBP1 encephalopathy (STXBP1-E), a developmental and epileptic encephalopathy that often begins in infancy. Most (75 – 89%) have seizures, typically starting in the first year (89%). Intellectual disability is common (90%), most severe or profound (64%). Half do not walk. Other common features include autism (19 – 42%), low tone, movement disorders, abnormal EEGs (> 60% with focal or multifocal epileptiform discharges), and abnormal MRI brain imaging (atrophy, thin corpus callosum, delayed myelination).^1-3^

SLC6A1 (solute carrier family 6 member 1) encodes GAT-1, a synaptic GABA reuptake transporter expressed in neurons and glia. Pathogenic mutations cause SLC6A1-related disorders (SLC6A1-RD), which begin in early childhood and are characterized by epilepsy (91%) and developmental delays (84%), typically in the mild to moderate range (64%). The epilepsy is typically generalized (seizure types like absence, atonic, myoclonic, generalized tonic-clonic), though it is sometimes focal (10%). Substantial minorities have an autism spectrum disorder (23%), movement disorder, or problems with attention or aggression.^4,5^

4-phenylbutyrate (4PB), formulated as sodium phenylbutyrate^6^ or glycerol phenylbutyrate,^7^ is an FDA-approved medication that reduces serum ammonia in people with urea cycle disorders. It can also improve symptoms of other disorders such as cholestasis,^8-11^ thalassemia,^12^ and maple syrup urine disease.^13^ 4PB crosses the blood-brain barrier (via the monocarboxylate transporter 1 mechanism)^14-16^ and has also been studied for neurological conditions such as spinal muscular atrophy,^17,18^ amyotrophic lateral sclerosis^19^, and Huntington’s disease.^20^

Preclinical studies suggested that 4PB may benefit people with STXBP1-E or SLC6A1-RD. In cellular and *C. elegans* models of STXBP1-E, 4PB rescued synaptic dysfunction via increased protein expression and prevention of misfolded protein aggregation.^21^ In cellular models of SLC6A1 dysfunction, 4PB reduced endoplasmic reticulum retention of the mutant protein, improved trafficking of the wild-type protein, and directly activated SLC6A1 (i.e., positive allosteric modulator of SLC6A1).^22,23^ In SLC6A1-mutant heterozygous mice, 4PB reduced spike-wave discharges.^22^

Here we report our initial experience treating STXBP1-E and SLC6A1-RD with 4PB, as glycerol phenylbutyrate. This report focuses on safety, tolerability, EEG, and seizure outcomes.

## METHODS

### Study Design and Ethics

We conducted a single-treatment group, multiple-dose, open-label pilot study of glycerol phenylbutyrate (brand name Ravicti; Amgen, Thousand Oaks, CA) in children with STXBP1-E and SLC6A1-RD at two centers (Weill Cornell Medicine, New York, NY, and Children’s Hospital Colorado, Aurora, CO). The institutional review board at both centers reviewed and approved the study. A data safety monitoring board met regularly to review the data and make recommendations. The trial is registered at clinicaltrials.org (NCT04937062). The United States Food and Drug Administration determined the study was exempt from an Investigational New Drug application (IND 168839).

### Enrollment Criteria

Each enrolled child was required to have a pathogenic (or likely pathogenic) mutation of STXBP1 or SLC6A1, a clinical picture consistent with the disorder, and age 15 or younger at enrollment. Before receiving the medication, the children were required to have normal liver and kidney function, normal platelets, normal QT interval, and no concurrent medical illness. We required an English-speaking caregiver to give consent. For STXBP1, children were required to have at least one seizure in the 4 weeks before enrollment. For SLC6A1, we planned to enroll some children without seizures (i.e., to develop early insight into potential effects on development independent of seizure control). Exclusion criteria included pregnancy, inborn errors of beta-oxidation, or use of alfentanil, quinidine, cyclosporine, or probenecid.

### Study Procedures

After enrollment, the study team met with the caregivers for at least four weekly visits (typically virtual) to review the history, gather prior medical records, and establish baseline characteristics and seizure frequency We collected seizure history at each visit through a structured questionnaire and caregiver interview.

The child was admitted for video EEG monitoring for three days and two nights. The first dose of glycerol phenylbutyrate (roughly 1/9^th^ of the goal dose) was given on the second day. After discharge, families were instructed to titrate glycerol phenylbutyrate over 1 – 2 weeks to a goal daily dose of 11.2mL / m2 (12.4 g/m2), maximum of 17.5 ml (19g), in three divided doses (i.e., the maximum dose in the FDA approved medication guide for glycerol phenylbutyrate).^24^ An investigator met with caregivers weekly via telemedicine to ask about seizure frequency and side effects. Site investigators were free to alter the titration schedule if there were side effects. The child was then admitted a second time for a single overnight video EEG, after at least six weeks of exposure to glycerol phenylbutyrate. EEG results are based on clinical EEG reports.

Caregivers who preferred to continue glycerol phenylbutyrate were enrolled in an extended use phase, and followed quarterly, including a yearly in-person visit. Otherwise, glycerol phenylbutyrate was weaned over two weeks.

Assessments of development, sleep, and quality of life were administered throughout the study and will be reported in detail separately.

The tables and text use the following terms for the subjects’ ages. A *neonate* is 28 days old or younger; an *infant* is older than 28 days but less than 2 years old; a *preschool age* child is at least 2 but less than 5 years old; a *school age* child is at least 5 but less than 12 years old; and an *adolescent* is at least 12 but younger than 18 years old.

## RESULTS

### Demographics

We enrolled 10 children with STXBP1 Encephalopathy and 10 with SLC6A1 Neurodevelopmental Disorder. At enrollment, the median age was 5 years (absolute range 4 months to 11 years). There were 14 boys and 6 girls. All were White; two were Hispanic.

### Baseline Function

All enrolled children were delayed in development. In the SLC6A1 group, nine of 10 had some spoken language; all could walk independently. In the STXBP1 group, none of the children had spoken language; 4 of 10 could walk independently. Two were infants at enrollment.

### Safety and Tolerability

There were six serious adverse events (SAEs), all unexpected. Of these six, one was attributed to 4PB – a preschool age girl with SLC6A1-RD was admitted for covid-19 and lethargy and found to have metabolic acidosis. 4PB was discontinued, and the acidosis resolved. Of note, this child re-enrolled into the extended-use arm after her seizures recurred a year later. (She takes sodium citrate to manage the acidosis.)

We asked for an independent review of one SAE – an infant with infantile epileptic spasms syndrome who was hospitalized for a recurrence of epileptic spasms shortly after starting 4PB and while still taking a low dose. The independent reviewer (a US-based board-certified child neurologist and board-certified epilepsy specialist) determined the recurrence was attributable to the natural history of STXBP1 disorders and that it was safe to continue the 4PB.

The other four SAEs included hospitalizations for (1) lethargy and thrombocytopenia (attributed to valproic acid toxicity), (2) ataxia (attributed to covid-19 infection), (3) respiratory distress (attributed to rhinovirus infection), and (4) polyuria and worsening seizure frequency (attributed to hyponatremia and oxcarbazepine toxicity).

Other common adverse events attributed to 4PB included a honey-like body odor (nearly all participants), sedation (8 participants), and anorexia (1 participant). These side effects often led to adjustment of the titration schedule and, for one child (SLC6A1), ongoing use of a substantially lower dose than recommended (3.3 ml per day instead of 8.5).

Of the 18 participants who remained in the study without interruption, 17 (94%) were taking a dose of 4PB within 10% of the target dose at the last clinical visit (after 2-3 years of participation). (Tables 1 and 2)

**Table 1.** Outcomes for children with STXBP1 developmental and epileptic encephalopathy.

| BASELINE |  |  |  | SEIZURE OUTCOMES |  |  | ANTI-SEIZURE MEDICATIONS |  |  | EEG FINDINGS (Clinical Reports) |  |  |  |
| --- | --- | --- | --- | --- | --- | --- | --- | --- | --- | --- | --- | --- | --- |
| Demographics* at enrollment | Mutation | Seizure Type(s) | Seizure Frequency | 10 week | 1 Year | Last Visit (2 - 3 years) | Enrollment | Last Visit | PBA (mL/day) Last Visit | Baseline (48 hours) | 6 weeks (24 hours) | 1 year (Routine) | 2 years (Routine) |
| *Short Term Seizure Freedom* |  |  |  |  |  |  |  |  |  |  |  |  |  |
| Preschool M, White, Not Hispanic | microdeletion (9q33.3-q34.13)¶ | Tonic | 13 of 35 days (15 seizures) | None†† | Monthly Seizures | Monthly Seizures | LEV, LTG, Keto | LTG | 9 (goal 8) | Normal | Discharges, Slow | Slow | Slow |
| Schoolage M, White, Not Hispanic | missense (R292P) | Tonic, electrographic | 2 of 28 days (2 seizures) | None | One breakthrough seizure | None | VPA | - | 13.5 (goal 14.5) | Sz, Discharges, Slow | Discharges, Slow | Slow | Slow |
| *Clinical Improvement* |  |  |  |  |  |  |  |  |  |  |  |  |  |
| Schoolage M, White, Not Hispanic | truncation (S311FfsX3) | Tonic | 15 of 63 days (17 seizures)† | 82% fewer days (3 of 70)<br>79% fewer seizures (4)† | Monthly Seizures, Clusters | Weekly-to-Monthly Seizures, Clusters | CBD, LTG | CBD, LTG | 9 | Sz, Discharges, Slow | Sz, Discharges, Slow | Discharges, Slow | Discharges, Slow |
| Schoolage F, White, Not Hispanic | truncation (K196X) | Tonic-clonic, tonic, epileptic spasms | 9 of 42 days (30+ seizures) | 60% fewer days (6 of 70)<br>76% fewer seizures (12) | Monthly Seizures, Clusters | Monthly Seizures, Clusters | LCM, OXC | - | 10.5 | Discharges, Slow | Ntch DIta, Slow | Ntch DIta, Slow |  |
| Schoolage M, White, Not Hispanic | missense (R190W) | Focal, Generalized convulsions | 10 of 42 days (22 seizures) | 52% fewer days (8 of 70)<br>62% fewer seizures (14) | Weekly-to-Monthly Seizures, Clusters | Weekly-to-Monthly Seizures, Clusters | OXC, CLB | LCM, CLB | 13.5 (goal 14.5) | Discharges, Slow | Slow | Slow | Slow |
| Schoolage F, White, Not Hispanic | missense (E283K) | Tonic, electrographic | 31 of 56 days (67 seizures) | 26% fewer days (20 of 49)<br>32% fewer seizures (40) | Weekly seizures | 8 month seizure free | CZP, ZNS, OXC | CZP, OXC, CBD, LCM, RUF | 10.5 | Sz, Discharges, Slow | Discharges, Slow | Discharges, Slow |  |
| *Indeterminate* |  |  |  |  |  |  |  |  |  |  |  |  |  |
| Infant F, White, Not Hispanic | missense (R406C) | Epileptic Spasms | None. (Resolved spasms) | Relapse of spasms, resolved‡ | None | None | LEV, ACTH | LEV, Keto | 6 | Discharges, Slow | Normal‡ | Slow | Slow |
| Preschool M, White, Not Hispanic | truncation (Q229X) | Epileptic Spasms | Daily clusters of epileptic spasms | Fewer spasms though still daily, less rescue medication | Daily seizures | Daily seizures | LEV, CBD, RUF, OXC | RUF, PER | 7.5 (goal 6.5) | Sz, Discharges, Slow | Sz, Discharges, Slow | Sz, Discharges, Slow |  |
| *Non-Responders* |  |  |  |  |  |  |  |  |  |  |  |  |  |
| Infant M, White, Not Hispanic | missense (R406C) | Epileptic Spasms | Daily clusters of epileptic spasms | Daily spasms (no change) | Frequent relapses of epileptic spasms | Frequent relapses of epileptic spasms | VGB, OXC | RUF, VGB, CBD | 7.5 | Sz, Discharges, Slow | Discharges, Slow | Discharges, Slow |  |
| Schoolage M, White, Not Hispanic | missense (P139L) | Tonic | 6 of 26 days (7 seizures) | 41% more days (15 of 46)<br>70% more seizures (21) | - | - | VGB, CBD | VGB, CBD | off | Sz, Discharges, Slow | Discharges, Slow |  |  |

\* Age ranges: Neonate = 0 to 28 days, Infant = > 28 days to < 2 years, Preschool = 2 years to < 5 years, Schoolage = 5 years to < 12 years, Adolescent = 12 years to < 18 years
† Excluding seizure cluster during a two-week period that included travel to New York for 1st admission (7 days with a seizure during these 14 days, 13 total seizures, 9 before first PBA dose, 4 after)
‡ Admitted urgently prior to the 6 week EEG for return of spasms. EEG at that time showed hypsarrhythmia and epileptic spasms. A course of steroids and increase dose of vigabatrin lead to resolution of the epileptic spasms, and the subsequent EEG was normal.
¶ Microdeletion includes the entire coding region of STXBP1
†† One seizure during up-titration to goal dose
PBA, glycerol phenylbutyrate; ACTH, adrenocorticotropic hormone; CBD, cannabidiol; CLB, clobazam; CZP, clobazepam; Keto, ketogenic diet; LCM, lacosamide; LEV, levetiracetam; LTG, lamotrigine; OXC, oxcarbazepine; PER, perampanel; RUF, rufinamide; VGB, vigabatrin; ZNS, zonisamide.
Sz, Seizures on EEG; Discharges, Epileptiform discharges on EEG; Slow, Slowing on EEG; Ntch DIta, Notched Delta

**Table 2.**
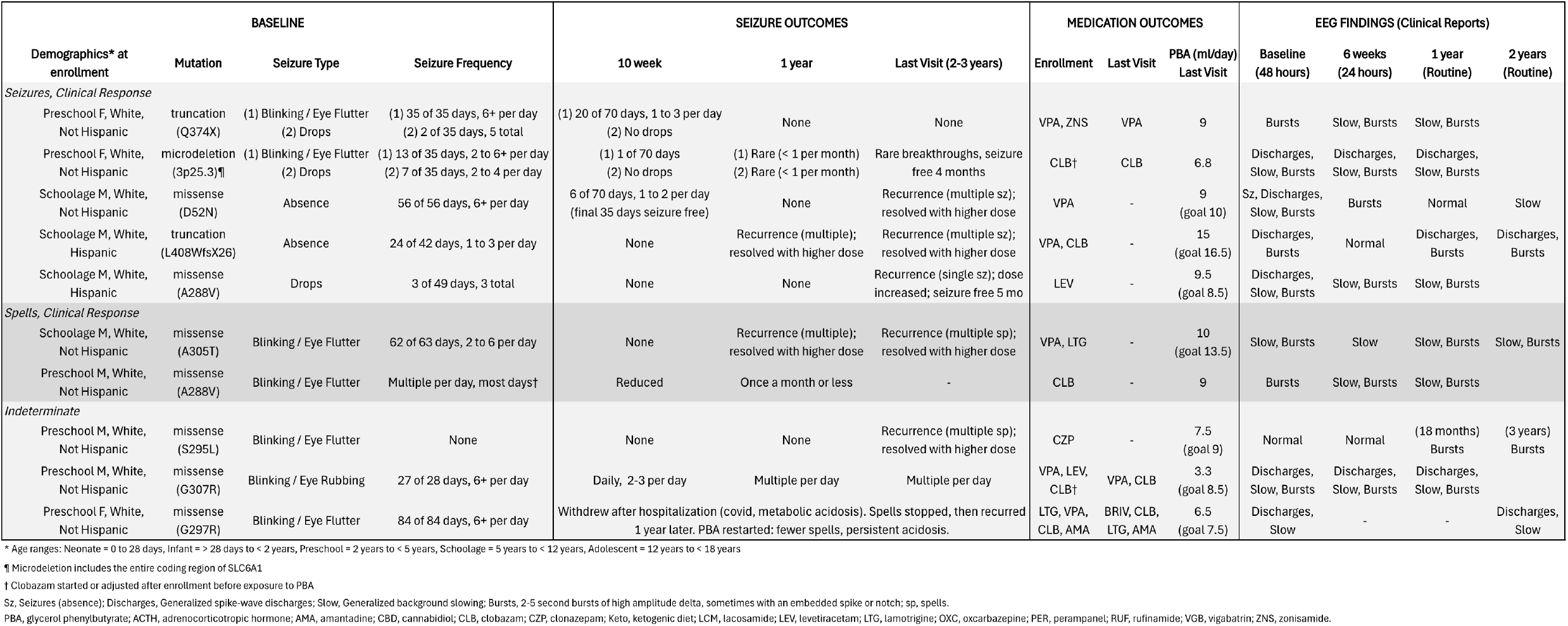
Outcomes for children with SLC6A1 neurodevelopmental disorder.

### STXBP1 Seizure and EEG Outcomes (Table 1)

In the STXBP1 group, seizure types included tonic seizures (6 participants), epileptic spasms (4), generalized convulsions (1), focal (1), and electrographic-only (2). In the initial observation period, two children had daily clusters of epileptic spasms, seven had daily to weekly seizures, and one child had no seizures (epileptic spasms resolved just before enrollment).

Six had a positive clinical response after the second admission (i.e., after 10 weeks of exposure): two were seizure-free after reaching the goal dose of 4PB, and four had reductions in seizures (26 – 82% fewer days with seizures, 32 – 79% fewer total seizures). Two had an indeterminate response -- one had a relapse of epileptic spasms at a low dose of 4PB, which then resolved with steroids and vigabatrin, and one continued to have daily epileptic spasms, though the mother reported fewer per day and less need for benzodiazepine rescue medication. There were two non-responders: one with persistent daily epileptic spasms and one with an increase in seizure frequency.

Responders included children with microdeletions, truncation, and missense variants. Of the three children with ongoing epileptic spasms as the primary seizure type during the observation period, none were responders.

Nine continued 4PB into the extended use period. At one year, eight of these nine had at least one seizure. At the last visit (2-3 years after enrollment), seven were still having seizures, ranging from monthly to daily. For four of these seven, the seizures tended to cluster. Two had weaned off other antiseizure medications and were taking only 4PB. The remaining seven continued antiseizure medications.

All 10 had abnormal EEGs. Abnormalities included generalized background slowing (10), epileptiform discharges (10), a notched delta pattern (1), and seizures (6). The captured seizures were subclinical (1), tonic (2), and epileptic spasms (3). There was some overall improvement in the EEGs over time. Of the six children with seizures on the initial overnight EEG, only two had seizures on the follow-up overnight EEG. At the last routine EEG, four no longer had epileptiform discharges.

### SLC6A1 Seizure and EEG Outcomes (Table 2)

In the SLC6A1 group, seizure types included absence seizures (2 participants) and “drops” (likely atonic or myoclonic-atonic seizures) (3). Seven children also had spells of blinking, eye fluttering, or eye rubbing, which did not have clear EEG correlates.

Among the nine children who returned for a second admission, seven had a positive clinical response on seizures (5 participants) or spells (2) after the second admission (10 weeks of exposure). Of these seven, three were free of events and four had more than 90% reduction. Responders included children with microdeletions, truncation, and missense variants.

For all seven, the clinical benefit continued through the last visit, 1-3 years after enrollment. One has been seizure-free with no recurrences. Five had occasional breakthrough seizures that resolved with increasing the dose of 4PB (i.e., weight adjustment). One had breakthrough seizures once a month or less.

We categorized three children as having an “indeterminate” response. One child had no seizures at baseline. This child developed blinking spells 2 years after enrollment, which resolved after increasing the dose of 4PB (i.e., weight adjustment). One reported ongoing daily blinking/eye rubbing spells but fewer per day after 10 weeks of exposure to 4PB (from 6+ per day to 2 or 3) – this child was also unable to tolerate the goal dose of the 4PB. We also categorized as “indeterminate” the child who disenrolled for metabolic acidosis and reenrolled for seizure control a year later – she had a clinical reduction in her spells after restarting 4PB, though she did not tolerate the goal dose due to persistent metabolic acidosis despite the use of sodium citrate.

At the last visit (2-3 years after enrollment) six of the ten children were no longer taking anti-seizure medications (other than 4PB). Two had reduced medication burden, and two were still taking the same number of anti-seizure medications.

All 10 had at least one abnormal EEG. Common EEG abnormalities included bursts of high-amplitude rhythmic generalized delta activity (9 participants), epileptiform discharges (6), and background slowing (7). For 1 participant, spike-wave discharges correlated with parental observations of eye fluttering, and we thus characterized those spells as absence seizures. For other participants, there was no clear correlation between EEG abnormalities and eye flutter events marked by caregivers.

There was no clear pattern of EEG evolution over the study. Three examples follow. For one child, the delta bursts faded after initial exposure to 4PB and did not return; for a second, the bursts faded then returned in subsequent EEGs; and for a third, the EEG was initially normal, then bursts developed 18 months after taking 4PB.

## DISCUSSION

### Summary of Findings

In this open-label study, glycerol phenylbutyrate improved seizure control in children with refractory epilepsy due to pathogenic mutations in STXBP1 (6 responders of 10) and SLC6A1 (7 responders of 10). This response rate (i.e., 60 – 70%) is higher than the placebo response rate in epilepsy drug trials, which ranges from 4 – 32%.^25,26^ There were no new safety concerns other than those already listed on the FDA-approved label. Metabolic acidosis was the side effect with the most serious consequences, leading to one participant’s withdrawal from the study. Despite the side effects, 90% (18 of 20) continued to take 4PB within 10% of the goal dose at the last visit.

### Repurposed Drugs

4PB for epilepsy is an example of a repurposed drug, i.e., a drug initially approved for one indication that may be efficacious for a different indication. Several FDA-approved drugs for epilepsy were initially approved for something else, such as acetazolamide (diuretic), fenfluramine (weight loss), and everolimus (renal cell cancer carcinoma). In the case of 4PB, no regulatory agency has approved its use for epilepsy, and thus its use is “off-label”. Regulations for off-label prescribing vary by jurisdiction. In the United States, clinicians may prescribe 4PB for epilepsy based on clinical judgment without regulatory approval,^27^ though the cost may be prohibitive. In Europe, regulations vary by country and are often more restrictive than in the United States.^27^

### Clinical Research Implications

We selected STXBP1 and SLC6A1 based on preclinical data known to our team at study inception.^21,22^ A growing body of preclinical data suggests the potential for 4PB to improve outcomes for other genetic epilepsies, including LGI1,^28^ GABA receptor mutations,^29^ creatine transporter disorder (SLC6A8),^30^ and SCN1A.^31^ Additional clinical trials in these disorders may broaden the pool of individuals who might benefit.

### Mechanisms

For genetic disorders with heterozygous mutations like STXBP1 and SLC6A1, there are multiple potential explanations for the action of 4PB, such as direct rescue of mutant proteins, enhancement of wild-type protein, beneficial epigenetic effects (e.g., histone deacetylase inhibition), or mitigation of toxic effects of misfolded proteins.^32^ Specific mechanisms of 4PB described in preclinical epilepsy models include protein stabilization (LGI1, STXBP1),^21,28^ improved trafficking (SLC6A1, SLC6A8, GABRG2),^21,22,30^ positive allosteric modulation (SLC6A1),^23^ prevention of misfolded protein aggregation (STXBP1, GABRG2),^21,29^ and reduction of endoplasmic reticulum stress and the unfolded protein response (GABRG2).^29^ The response of two fully haploinsufficient children (i.e., microdeletions, one in each group), suggests that improved wild-type function is an important mechanism of 4PB. However, multiple actions may be responsible for the observed clinical improvements. Additional bench neuroscience to understand the mechanisms of 4PB may help predict which other populations of people with epilepsy would benefit from the drug.

### Biomarkers

EEG patterns evolved for several children throughout the study, with reduction of discharges in some children with STXBP1, and a more complex evolution of bursts in children with SLC6A1. It is unclear if these changes represent the natural history of EEG patterns associated with the disease or the waxing-waning effectiveness of 4PB or other anti-seizure medications. Additional work would be valuable to understand if there are EEG features that might be used as a biomarker for disease activity and/or disease progression, like delta power in Angelman Syndrome.^33,34^

### Limitations

Open-label studies are prone to multiple sources of bias due to the absence of blinding, the lack of a control group, and no randomization. Selection bias is also likely, as we preferentially enrolled children with greater seizure burdens. We did not recruit a racially and ethnically diverse cohort, with no Black participants and only 10% Hispanic (versus 19% of the US population). This is a well-described limitation throughout clinical research, limiting generalizability.^35^ Several of our assessments were not standardized – for example, physicians who were study investigators performed the clinical interpretation of some EEGs. Seizure counts are subject to bias.^36^ Staring spells and absence seizures (i.e., in the SLC6A1 group), are difficult to count reliably. Tonic seizures (i.e., in the STXBP1 group) often happen at night and can be undercounted.

### Conclusions

For children with STXBP1 and SLC6A1 disorders, glycerol phenylbutyrate was well tolerated with no new safety concerns. The finding of seizure reduction is promising and should be confirmed through additional clinical trials.

## Data Availability

All data produced in the present study are available upon reasonable request to the authors.

## ACKNOWLEDGEMENTS

ZG, AS, and NB had full access to all the data in the study and take responsibility for the integrity of the data and the accuracy of the data analysis. A copy of the data is available on request to the corresponding author (ZG), with an approved proposal and signed data use agreement. We did not use artificial intelligence or large language models in the writing of this manuscript, other than via commercial software to check spelling and grammar (Grammarly, Inc. San Francisco, CA). We are grateful to the parents and caregivers who enrolled their children into this study – their insights and observations contributed greatly to our understanding of the findings.

This investigator-initiated study was funded by multiple sources, including for-profit pharmaceutical companies (Horizon Therapeutics, now Amgen), non-profit caregiver-led advocacy groups (STXBP1 Foundation, SLC6A1 Connect), academic medical centers (Weill Cornell Medicine and Penn Medicine via the Orphan Disease Center), and private philanthropy (Morris and Alma Schapiro Fund, Clara Inspired).

Dr. Grinspan (corresponding author) currently receives research funding from Weill Cornell Medicine, NIH/NINDS (R01NS130113), Amgen, Harmony Biosciences, SLC6A1 Connect, STXBP1 Foundation, the Morris and Alma Schapiro Fund, the Jain Foundation, and the D’Addario Foundation. Dr. Grinspan has conducted paid consulting work for Capsida Therapeutics, Mahzi Therapeutics, Encoded Therapeutics, and Neurvati Neurosciences.

Dr. Burré receives research funding from NIH/NINDS (R01NS113960, R01NS121077, R01AG083949, RF1NS126342, R01NS113960), and is a member of the scientific advisory board for the SNAP-25 foundation and the VAMP2 foundation.

Dr. Cross receives research support from Amgen, STXBP1 Foundation, and SLC6A1 Connect.

Dr. Ross receives research support from the NIH (R01NS105477, U54NS117170, OT2OD037643).

Dr. Kang receives research support from NIH/NINDS (R01NS121718) and SLC6A1 Connect.

Dr. Miele consults for Biogen Inc., Capsida Biotherapeutics, and Encoded Therapeutics. She receives research funding from the STXBP1 Foundation.

Dr. Demarest has consulted for Biomarin, Neurogene, Marinus, Tysha, Ultragenyx, UCB, Capsida, Encoded, Longboard, Mahzi Therapeutics, and Ovid Therapeutics. He has funding from the NIH, Project 8P, and Mila’s Miracle Foundation. He also serves on advisory boards for the non-profit foundations Rare X, SLC6A1 Connect, Project 8P, Ring14 USA, FamilieSCN2A, and N of 1 Collaborative.

